# Timing and risk of age-related conditions in individuals with Down syndrome

**DOI:** 10.64898/2026.09.21.26363043

**Authors:** Hannah F. Tavalire, Akaninyene Noah, Veronica Fitzpatrick, Hannah Graham, Shelly Verma, Laura Krohn, Victoria L. Fleming-Batayneh, Katie Frank, Daniel Vockeroth, Brian Chicoine

## Abstract

Individuals with Down syndrome (DS) exhibit accelerated biological aging and unique patterns in disease prevalence. However, empirical characterizations of relative risk and age at onset of disease in this population are not common. Here we aimed to quantify the difference in risk and timing of age-related conditions between individuals with DS when compared to typically developing (TD) matched controls. We used semiparametric Cox proportional hazards models and Restricted Mean Event-Free Age (RMEA) to estimate differences in risk and approximate average age at onset of cataracts, cognitive decline, death, menopause, and osteoporosis. Individuals with DS were at higher risk of all age-related conditions or events and experienced these events at younger ages, excluding menopause. We observed a 24.5-fold higher risk of cognitive decline in individuals with DS (p<0.001), and between a 2- and 4-fold higher risk of the other age-related events or conditions (p<0.01). Cognitive decline and death occurred more than 14 years earlier in individuals with DS and were more likely to co-occur (p<0.001) compared to in TD individuals. These findings have important implications for tailored screening and prevention efforts in aging individuals with DS, as this population experiences a distinct cadence of age-related events.

**Author Summary:** Here we characterize differences in when individuals with Down syndrome (DS) experience age-related conditions and events relative to typically developing (TD) individuals. Individuals with DS experienced cataracts, cognitive decline, death, and osteoporosis at a younger age and were at higher risk for these conditions than typically developing individuals. Most notably, we found that individuals with DS are at a 24.5-fold greater risk of cognitive decline and experience cognitive decline 14.7 years before TD individuals. We observed a similar age disparity in time of death (14.2-year difference), with median age at time of death estimating an even more substantial gap of 23.4 years between cohorts. We detected no difference in the timing or risk of menopause between groups. We also observed that the rate at which individuals with DS experience more than one of these age-related conditions differed significantly from TD controls, with death and cognitive decline co-occurring most often. These findings exemplify the need for tailored screening and prevention efforts in individuals with DS, moving away from general guidelines developed in TD populations.

## Introduction

Down syndrome (DS) is the most common chromosomal abnormality among live-born infants in the United States, affecting 1 in 700 live births and continuing to increase in prevalence(1, 2). The life expectancy of individuals with DS has more than doubled to approximately 60-65 years over the past 40 years but remains much lower than in typically developing (TD) individuals (2-4). Given this substantial progress in life expectancy, the number of adults with DS has increased 8-fold since 1950 (5). With a rapidly growing aging population of individuals with DS, we are faced with the welcome challenge of developing tailored care guidelines for this population. Though recent efforts have turned toward specialty care protocols for individuals with DS, available research often leaves workgroups with a limited evidence base, leading to few conclusive recommendations (6). In the absence of widely available tailored guidance, clinicians currently follow established age-based standards for screening and prevention, rooted in onset timing and disease risk observed in TD individuals. It remains unknown what critical care windows may be missed for patients with DS under this care model.

Individuals with DS not only have a shorter average lifespan, but exhibit accelerated biological aging relative to TD individuals (7). Additionally, conditions like obstructive sleep apnea, autoimmune disorders, and Alzheimer’s disease occur at much higher prevalence in individuals with DS (8, 9). In contrast, some illnesses are less common in individuals with DS (e.g., solid tumors (10) and hypertension (11)). Given this mix of severity and timing of disease risk, and the apparent disconnect in aging progression relative to TD individuals, defining age-related risk of disease in individuals with DS has no unidirectional solution. Simply reducing the age at which related screening and prevention efforts begin would likely lead to unnecessary and invasive medical procedures in a population already burdened by distrust of the medical community (12). Characterizations of lifespan and disease prevalence in individuals with DS have emerged in recent years, laying the foundational understanding of disease occurrence and cause of death in this population (9, 13). More recent cohort comparison studies have also begun to characterize differences in lifespan and the prevalence of conditions in individuals with DS relative to matched controls (14-17). However, explicit comparisons of age at disease onset are often limited to focused investigations of single conditions, such as dementia (e.g., (18-21); however, see Baksh *et al*., (22) for a recent, comorbidity-focused approach). Here we utilize semiparametric time to event modeling and Restricted Mean Event-Free Age (RMEA) as a two-pronged approach to (1) characterize heterogeneity in disease risk and (2) approximate timing of onset of conditions and events commonly associated with aging. We observed stark differences in disease risk and timing brought on by disparate patterns in biological aging between individuals with and without DS.

## Results

Individuals with DS were matched 10:1 to TD controls as described in the methods section. Demographics of each cohort are described in Table 1. Briefly, we observed similar mean ages and a slight male excess in both cohorts. Individuals with DS had, on average, twice the number of total encounters compared to matched TD individuals. Race/ethnicity composition was acceptably balanced. Most individuals across cohorts were covered under commercial insurance; however, individuals with DS were more likely to have Medicare or Medicaid insurance than TD individuals.

**Table 1:**
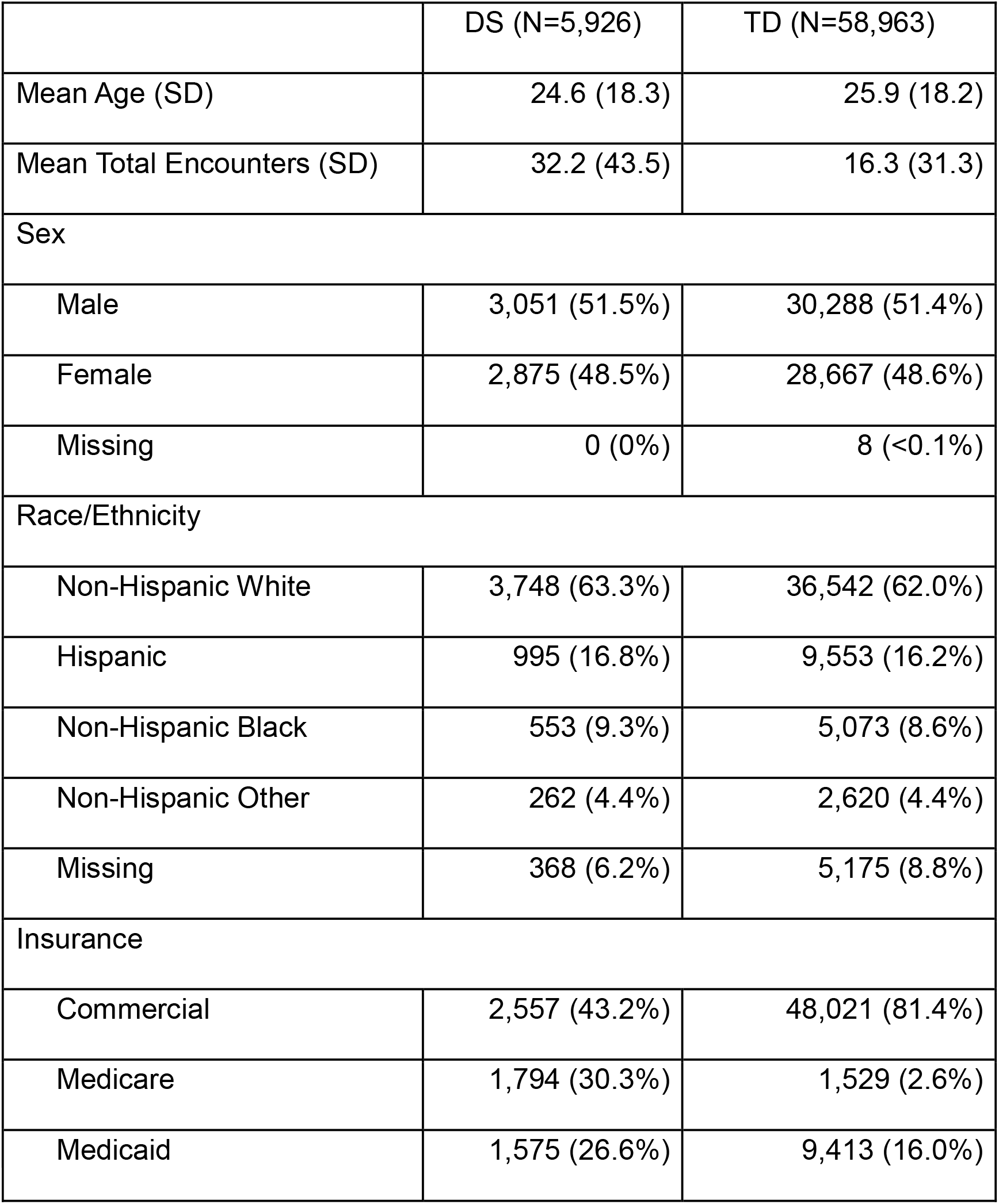
Cohort Demographics and Baseline Characteristics.

|  | DS (N=5,926) | TD (N=58,963) |
| --- | --- | --- |
| Mean Age (SD) | 24.6 (18.3) | 25.9 (18.2) |
| Mean Total Encounters (SD) | 32.2 (43.5) | 16.3 (31.3) |
| Sex |  |  |
| Male | 3,051 (51.5%) | 30,288 (51.4%) |
| Female | 2,875 (48.5%) | 28,667 (48.6%) |
| Missing | 0 (0%) | 8 (<0.1%) |
| Race/Ethnicity |  |  |
| Non-Hispanic White | 3,748 (63.3%) | 36,542 (62.0%) |
| Hispanic | 995 (16.8%) | 9,553 (16.2%) |
| Non-Hispanic Black | 553 (9.3%) | 5,073 (8.6%) |
| Non-Hispanic Other | 262 (4.4%) | 2,620 (4.4%) |
| Missing | 368 (6.2%) | 5,175 (8.8%) |
| Insurance |  |  |
| Commercial | 2,557 (43.2%) | 48,021 (81.4%) |
| Medicare | 1,794 (30.3%) | 1,529 (2.6%) |
| Medicaid | 1,575 (26.6%) | 9,413 (16.0%) |

### Relative risk and timing of age-related conditions

Overall, individuals with DS were at higher risk for all age-related conditions or events (Fig 1) and experienced these conditions or events at higher prevalence and at younger ages than TD individuals, excluding menopause (Fig 2; Table 2). Individuals with DS were diagnosed with cataracts on average 3.6 years earlier than TD individuals and had an adjusted 4-fold greater risk (CI[3.17, 5.05], p<0.001; Table 2A, Figs 1A & 2A). Cognitive decline showed the largest age gap and disparity in risk, with individuals with DS experiencing cognitive decline an average of 14.7 years before their TD counterparts, with an adjusted 24.5-fold risk (CI[20.5, 29.4], p <0.001; Table 2B, Figs 1B & 2B). Death showed a similar age gap between cohorts, with all-cause mortality occurring in individuals with DS 14.2 years earlier, on average, with median age at time of death estimating an even more substantial gap of 23.4 years between cohorts. Individuals with DS also had a 2.0-fold higher adjusted risk of death (CI[1.3, 3.1], p = 0.002; Table 2C, Figs 1C & 2C) and accrued an addition 2.7% increase in risk for each year of age (CI[1.02, 1.04], p < 0.001). Menopause was observed at similar frequency between cohorts, and no significant differences were observed in risk (CI[0.70, 1.18], p = 0.44) or timing of this event (Table 2D, Figs 1D & 2D). Individuals with DS were at a 3.8-fold higher risk of osteoporosis (CI[3.82, 5.09], p < 0.001; Table 2E, Figs 1E & 2E) compared to TD individuals and were diagnosed with osteoporosis 3.2 years earlier on average. Observed frequencies and age-ranges for each event are reported in Table 2. Eighty years was selected as the time horizon (tau) for RMEA estimation, as this was the last decade in which both cohorts had several individuals still alive. Full risk model results including covariates are presented in Supplementary Table 1.

**Table 2:** Time to Event Model Results and RMEA/RMST Estimates.

|  | DS (N=5,926) | TD (N=58,963) |
| --- | --- | --- |
| **A. CATARACTS** |  |  |
| Observed Events (%) | 139 (2.4%) | 312 (0.5%) |
| Age Range at Time of Event | 16.3 – 74.9 | 18.2 – 87.8 |
| Cox Hazard Ratio | 4.0** | *ref* |
| Restricted Mean Event-Free Age | 74.4 [73.2, 75.7] | 78.0 [77.7, 78.3] |

| **B. COGNITIVE DECLINE** |  |  |
| --- | --- | --- |
| Observed Events (%) | 502 (8.5%) | 209 (0.4%) |
| Age Range at Time of Event | 15.1 – 72.1 | 35.4 – 87.8 |
| Cox Hazard Ratio | 24.5** | *ref* |
| Restricted Mean Event-Free Age | 63.9 [63.0, 64.8] | 78.6 [78.4, 78.8] |
| **C. DEATH** |  |  |
| Observed Events (%) | 545 (9.2%) | 615 (1.0%) |
| Age Range at Time of Event | 0 - 87.5 | 0 - 98.8 |
| Cox Hazard Ratio | 2.0* | *ref* |
| Restricted Mean Survival Time | 63.1 [62.4, 63.8] | 77.3 [77.0, 77.6] |
| Median Age at Time of Event | 57.7 [56.8, 58.7] | 81.1 [78.7, 83.8] |
| **D. MENOPAUSE** |  |  |
| Observed Events (%) | 72 (2.5%) | 713 (2.4%) |
| Age Range at Time of Event | 23.8 – 64.5 | 21.8 – 91.7 |
| Cox Hazard Ratio | 0.9 | *ref* |
| Restricted Mean Event-Free Age | 76.0 [75.0, 76.9] | 75.0 [74.6, 75.4] |
| **E. OSTEOPOROSIS** |  |  |
| Observed Events (%) | 81 (1.4%) | 222 (0.4%) |
| Age Range at Time of Event | 21.2 – 80.4 | 7.4 – 88.1 |
| Cox Hazard Ratio | 3.8** | *ref* |
| Restricted Mean Event-Free Age | 75.5 [74.0, 76.9] | 78.7 [78.5, 78.9] |

\*p<0.01, \*\*p<0.001, 95% CIs in brackets where applicable.
Individuals with missing covariate data were excluded from time to event models only.

**Fig 1:**
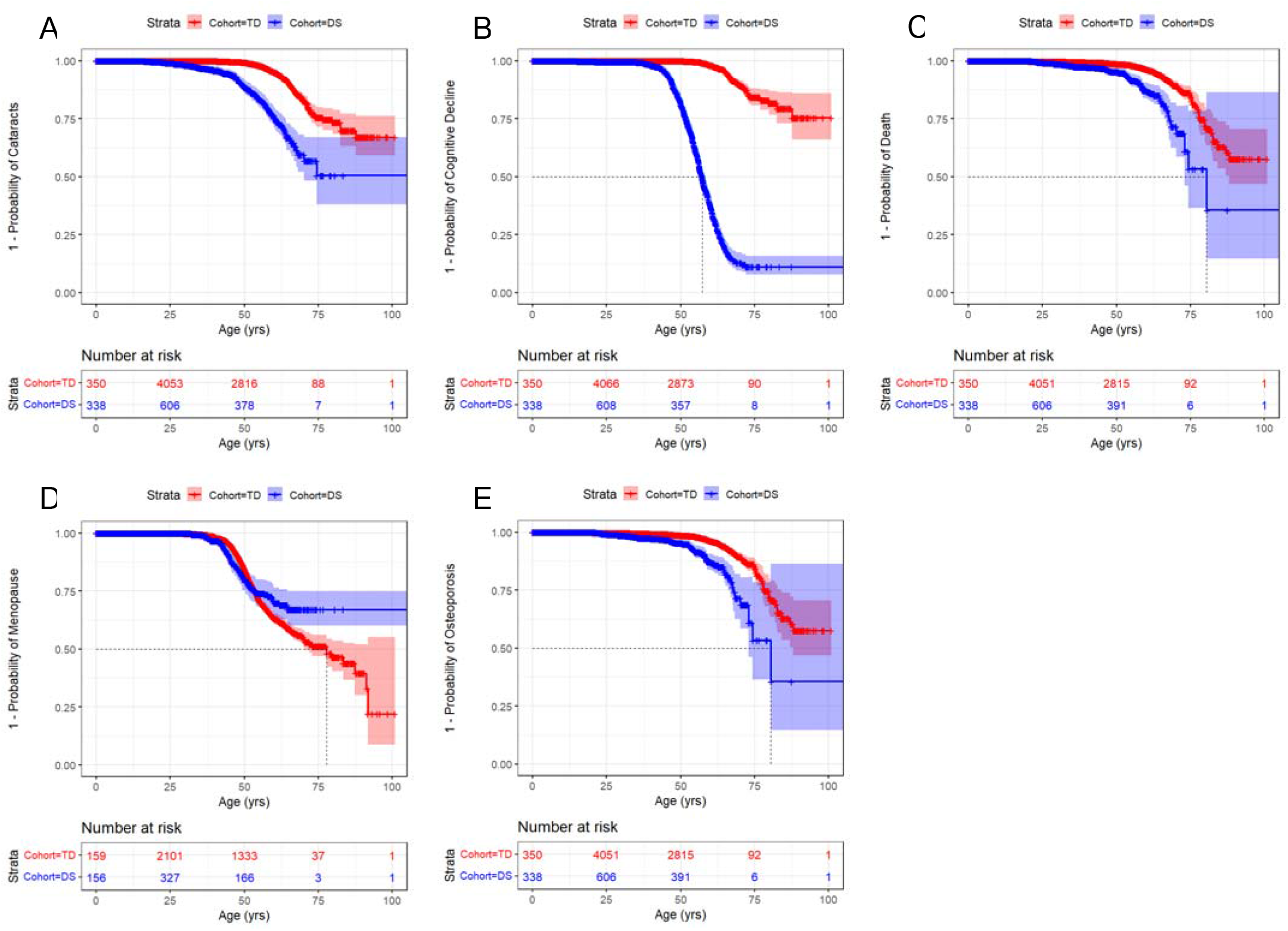
Kaplan-Meier time-to-event curves for each condition of interest as patients age. Time to event curves for individuals with Down syndrome (DS) are in blue, while those of typically developing (TD) individuals are in red. Curves depict the probability of each event not occurring over time (1 – p). Thus, a steeper slope implies earlier age at onset and a more frequently experienced event. Number of individuals at risk for each cohort are detailed along the bottom of each plot in 25-year intervals and reflect age-specific sample sizes due to interval censoring.

**Fig 2:**
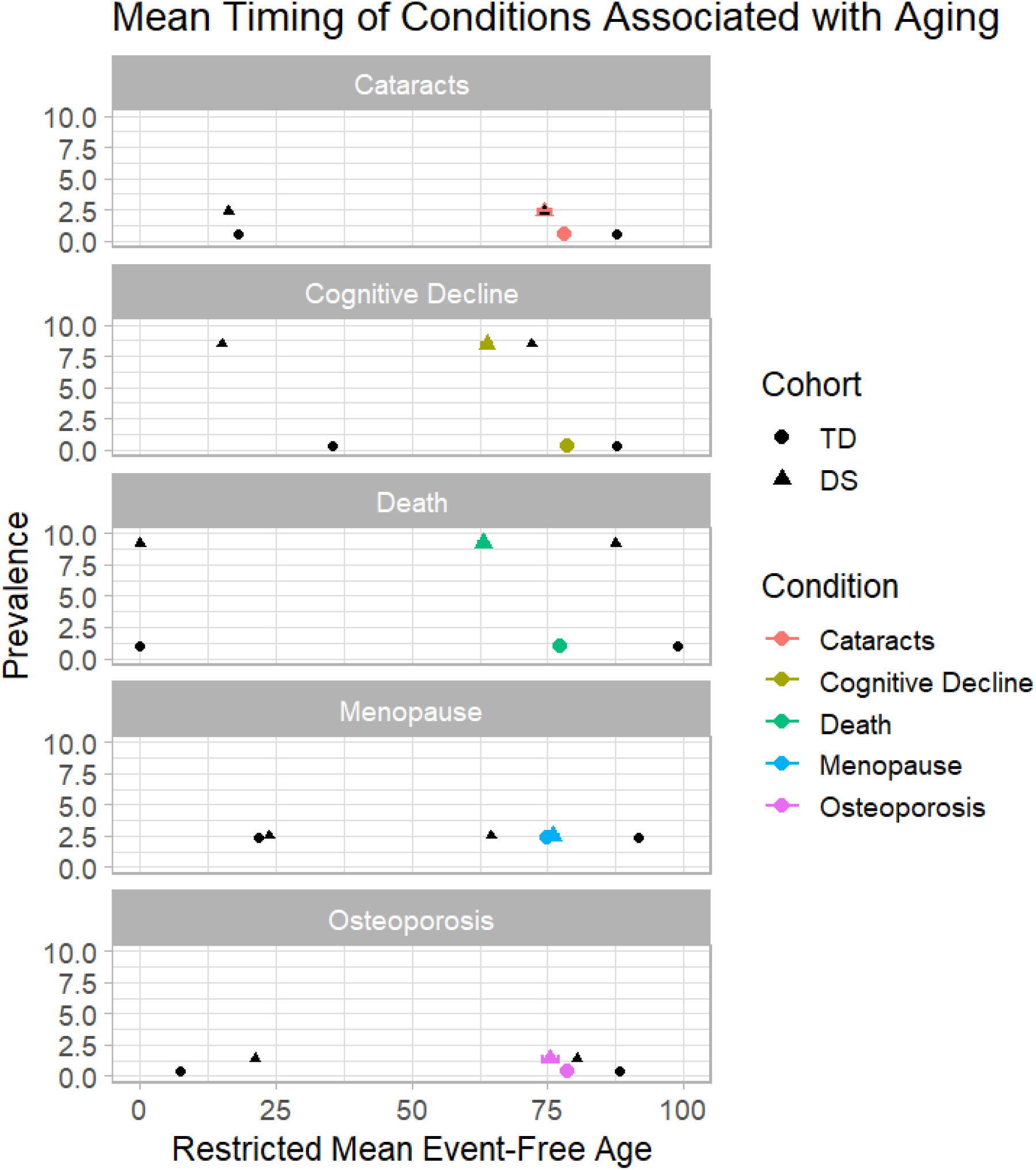
Restricted Mean Event-free Age (RMEA) 129 for each condition or event of interest. Estimated RMEAs are presented for individuals with Down syndrome (DS, triangles) and typically developing individuals (TD, circles), color coded by each condition/event. Minimum and maximum observed values are denoted in black for each cohort and condition. Cohort level prevalence is indicated on the y-axis. RMEA estimates may lie outside of the observed min-max range due to censoring of older individuals that had no observed event.

### Cooccurrence of age-related conditions

Observed cooccurrence of age-related conditions differed significantly between individuals with DS and TD individuals (X^2^_df=9_ = 216.9; p < 0.001; Table 3). When comparing observed to expected counts for each condition cooccurrence pair, under the null hypothesis that cooccurrence frequencies did not differ between the cohorts, we observed several notable differences in observed frequencies. In individuals with DS, cognitive decline and death were observed to cooccur at a much higher rate than expected (4.1%) while events cooccurring with menopause were generally under-observed (<1%). In TD individuals, we observed an excess cooccurrence of menopause with all other conditions, though absolute frequencies of these combined events remained low. The combination of death and cognitive decline occurred at a much lower rate than expected (<0.01%) in TD individuals, suggesting a disparity in survivorship following cognitive decline in individuals with DS relative to TD individuals.

**Table 3:** Cooccurrence Matrix of Age-Related Events in Individuals with DS (lower triangle) and TD Individuals (upper triangle).

| DS (%) \ TD (%) | Cataracts | Cognitive Decline | Death | Menopause* | Osteoporosis |
| --- | --- | --- | --- | --- | --- |
| Cataracts | 312 (0.5%)<br>139 (2.3%) | 12 (<0.1%) | 20 (<0.1%) | 39 (0.1%) | 19 (<0.1%) |
| Cognitive Decline | 48 (0.8%) | 209 (0.4%)<br>502 (8.5%) | 31 (<0.1%) | 15 (<0.1%) | 10 (<0.1%) |
| Death | 47 (0.8%) | 240 (4.1%) | 615 (1.0%)<br>545 (9.2%) | 14 (<0.1%) | 20 (<0.1%) |
| Menopause* | 7 (0.2%) | 13 (0.5%) | 11 (0.4%) | 713 (1.2%)<br>72 (1.2%) | 36 (0.1%) |
| Osteoporosis | 7 (0.1%) | 25 (0.4%) | 35 (0.6%) | 1 (<0.1%) | 222 (0.4%)<br>81 (1.4%) |

Much Less
Somewhat Less
As Expected
Somewhat More
Much More
Total $N_{DS} = 5,926$ ; Total $N_{TD} = 58,963$
Values along the diagonal correspond to each single condition
Values below the diagonal are co-occurrence counts in individuals with DS (left justified), above the diagonal are for the TD cohort (right justified)
Percentages are % of total cohort with those cooccurring conditions

## 3. Discussion

We observed marked differences in age-related risk and approximate age of onset in age-related conditions and events in individuals with DS when compared to a TD control cohort, with most conditions and events occurring 5 to 20 years earlier in individuals with DS. Additionally, we observed significant differences in cooccurrence of age-related conditions and events between cohorts, with high frequency of cognitive decline cooccurring with death and cataracts, and lower frequency of any condition cooccurring with menopause in individuals with DS. Taken together, these findings have important implications for specialty care, as screening and prevention efforts anchored in general eldercare guidelines may miss critical earlier windows of risk or the unique cadence of comorbid onset of conditions in individuals with DS.

Excluding menopause, all evaluated conditions and events occurred earlier in individuals with DS. Interestingly, age at onset of cognitive decline and death were the most disparate *between* cohorts, spanning a ∼14 year difference between cohorts, but the relative timing of these events was most closely linked *within* a cohort, suggesting a distinct increase in risk of death following the onset of cognitive decline in individuals with DS compared to TD individuals. This finding is further corroborated in the co-occurrence analysis in which comorbid cognitive decline and death were observed at a higher frequency than expected in individuals with DS. Overall, individuals with DS were at a 24.5-fold greater risk of cognitive decline. Though increased prevalence of cognitive decline is well-established in individuals with DS (23-25), this paper is the first to our knowledge to quantify the relative risk of cognitive decline and other age-related conditions in a cohort study compared to matched controls.

We observed significant increases in relative risk of cataracts (4.0x), death (2.0x), and osteoporosis (3.8x) in individuals with DS, though at a less staggering difference than our observations of cognitive decline. Interestingly, though menopause was observed at the same frequency in both cohorts, conditions co-occurred with menopause less often than expected in individuals with DS (except osteoporosis), while menopause was observed to co-occur more frequently with other conditions than expected in the TD cohort. Additionally, the RMEA for menopause falls outside of the observed range of ages for this event in individuals with DS. Taken together, these observations are likely due to under-detection or under-reporting of menopause in women with DS, since menopause is a ubiquitous female experience and women with DS have been previously reported to experience menopause roughly 5 years earlier than TD women (26).

Disparities in the lifespan of individuals with DS present a significant methodological challenge in matched cohort comparison studies. For example, a 40-year-old with DS may - hypothetically – be biologically comparable to a 60-year-old without DS, complicating traditional age-based matching methods. Therefore, the assumption in matched cohort studies that the baseline age-specific risk of a certain condition is the same across cohorts, thus allowing any departures in risk to be attributed to the exposure (27), is likely violated when comparing individuals with DS to typically developing controls of the same age. Furthermore, TD individuals often live much longer than individuals with DS, making the comparison of age-matched individuals impossible past a certain age. Here we demonstrate differences in approximate disease timing between cohorts (i.e., different age at onset), adding to existing evidence that individuals with DS undergo accelerated biological aging. Future work will include refinement of matching procedures for this population to also assess *age-independent* relative risk. However, we find age-dependent assessments of risk to be most relevant to the design of specialty care guidelines for individuals with DS.

Most limitations of this study arise from the imperfect surveillance inherent to EMR data. We are only able to detect a diagnosis of interest if noted during an encounter, leading to potentially missed conditions and some uncertainty around timing of condition onset. However, we account for the bulk of this uncertainty in the current study by using time to event models fit with interval censoring. We also observed very low rates of menopause and death documentation in both cohorts, likely leading to global underestimation of the prevalence of these events. Previous work has reported individuals with DS experiencing menopause earlier than their TD counterparts (26), a pattern suggested by the time to event curve in Fig1D but not corroborated by formal analyses. Several barriers remain to accurate diagnoses in individuals with DS involving subjective assessments or standardized instruments validated in TD individuals, such as the evaluation of cognitive decline or bone density. It is well-established that diagnostic overshadowing can obscure the ability of non-specialist clinicians to make accurate assessments of age-related change in individuals with DS (28), leading to potential underdiagnosis or delays in diagnosis of conditions, especially those involving mental faculty. Furthermore, individuals with DS often have artificially low bone density test results due to their shorter average height (29, 30), complicating diagnosis of osteoporosis. These examples serve as further evidence that screening and prevention efforts require further tailoring for individuals with DS to promote equity in care as this population ages. Future directions will build upon this foundational work in relative risk and timing of age-related events in individuals with DS to ultimately inform specialty care guidelines tailored to this population.

## Materials and Methods

### Study setting and population

This study used retrospective health data from a large healthcare network in the Midwest United States (Advocate Health; AH). AH consists of 25 hospitals, over 500 clinical care locations, and the largest specialty care center for adults with DS in the US. The Institutional Review Board approved this study under exemption category 4 (IRB #00132715) and waived informed consent requirements, as the study involved only secondary use of identifiable information.

Individuals with DS were identified using the International Classification of Diseases, 9th and 10^th^ revisions, Clinical Modification (ICD-9/10-CM) codes of 758.0 and Q90, respectively (the data span periods when both revisions were in use). Data on encounters and diagnoses were extracted from the Electronic Medical Record (EMR) of all individuals with DS seen from January 2005 and May 2025. During this study period, we identified 5,926 individuals with DS. TD individuals were matched to the DS cohort at a ∼10:1 ratio based on age at first encounter, sex, race/ethnicity, year of first encounter, and encounter frequency during the year of first encounter, for a total of 58,963 TD controls.

### Data sources

All data were collected via retrospective EMR extract. Individuals with DS and TD individuals had a total of 553,604 and 2,248,202 encounters recorded in the EMR across the study period, respectively. We considered the following age-related conditions or events of interest: Cataract diagnosis, cognitive decline diagnosis, date of death, menopause diagnosis, and osteoporosis diagnosis. Any age-related condition of interest was assigned to the first encounter at which it was diagnosed by ICD-10 code in the EMR (see Supplementary Table 2 for ICD-10 codes included for each age-related condition group). Death dates were used when available to denote exact timing of death (available for 545 individuals with DS and 615 TD individuals), however death dates are not systematically recorded at AH if a patient passes away outside of an AH facility.

### Statistical analyses

Continuous variables are summarized by means and standard deviations by cohort. Count data are represented as absolute counts and percentages by cohort. Model and metric estimates are presented with 95% confidence intervals (CIs) and associated p-values, where appropriate. Statistical significance was assessed against a Bonferroni-corrected p-value of 0.01 to account for the five age-related outcomes considered here. All data manipulations and analyses were performed in R v4.5.1 (31).

#### Time to event modeling

We used two variations of the Cox Proportional Hazards model to estimate relative risk in age-related conditions between individuals with DS and TD individuals. Time to event modeling for age-related diagnoses (cataracts, cognitive decline, menopause, osteoporosis) was conducted using semi-parametric Cox Proportional Hazards models fit with interval censoring in the *icenReg* package (32). Interval censoring allows for uncertainty in the true onset timing of each condition while anchoring the surveillance end point to the date of diagnosis. The model for age at death was fit using a traditional Cox Proportional Hazards model fit in the *survival* package (33), since exact death dates were known. Individuals with less than 2 visits or who were missing covariate data were removed from the analysis dataset, yielding cohorts of 5,558 (DS) and 53,780 (TD) for time to event models. All Cox PH models were adjusted for sex (except menopause model), race/ethnicity, and year of AH system entry. Proportional hazards assumptions were tested and time-varying terms (death model) or stratification by sex (osteoporosis) were used to resolve any detected violations. If the median time to event was reached for a given model, it is reported (only for death).

#### Restricted Mean Event-Free Age (RMEA)

The evaluation of relative age at onset can be difficult when considering rare events, since a median time to event is often not reached in these models. To circumvent this common challenge, we used Restricted Mean Event-Free Age (RMEA; an extension of Restricted Mean Survival Time (RMST)) to estimate the average event-free years experienced by each cohort up to a horizon (tau) of 80 years. RMEA is highly interpretable as ‘life expectancy’ prior to an event, or the approximate average ‘age at onset,’ and performs well when assumptions of proportional hazards are not upheld (34, 35). Eighty years was selected as the time horizon for these analyses, as this was the last decade in which both cohorts had several individuals still alive. RMEAs were estimated using the *survRM2* package in R (36). RMEAs were considered significantly different if 95% CIs were non-overlapping between cohorts.

#### Age-related event cooccurrence

Age-related event cooccurrence was evaluated by counting the individuals in each cohort who experienced each pair of events. Significant difference in proportions experiencing each pair of events was tested between cohorts using a Pearson’s Chi-Squared Test. Observed counts for each event pair were compared to expected values to determine directionality of observed departure in frequency.

## Data Availability

All data produced in the present study are available upon reasonable request to the authors

## Acknowledgments

This work was supported by an Advocate Aurora Research Institute LiveWell intramural grant awarded to B. Chicoine. The authors would like to thank the Advocate Health research analytics teams for their assistance with data acquisition, extraction, and quality assurance. The authors would also like to thank the dedicated healthcare staff at Advocate Health involved in the collection, management, and maintenance of the EMR data used in this study. 4

## Supporting information captions

**Supplementary Table 1**. Full time-to-event model results for each outcome of interest.

**Supplementary Table 2**. ICD-10 codes used to identify cases of each condition of interest.

